# Investigating Blunt Force Trauma to the Larynx: The Role of Inferior-Superior Vocal Fold Displacement on Phonation

**DOI:** 10.1101/2020.11.18.20234203

**Authors:** Molly E. Stewart, Byron D. Erath

## Abstract

Blunt force trauma to the larynx, which may result from motor vehicle collisions, sports activities, etc., can cause significant damage, often leading to displaced fractures of the laryngeal cartilages, thereby disrupting vocal function. Current surgical interventions primarily focus on airway restoration to stabilize the patient, with restoration of vocal function usually being a secondary consideration. Due to laryngeal fracture, asymmetric vertical misalignment of the left or right vocal fold (VF) in the inferior-superior direction often occurs. This affects VF closure and can lead to a weak, breathy voice requiring increased vocal effort. It is unclear, however, how much vertical VF misalignment can be tolerated before voice quality degrades significantly. To address this need, the influence of inferior-superior VF displacement on phonation is investigated in 1.0 mm increments using synthetic, self-oscillating VF models in a physiologically-representative facility. Acoustic (SPL, frequency, H1-H2, jitter, and shimmer), kinematic (amplitude and phase differences), and aerodynamic parameters (flow rate and subglottal pressure) are investigated as a function of inferior-superior vertical displacement. Significant findings include that once the inferior-superior medial length of the VF is surpassed, sustained phonation degrades significantly, becoming severely pathological. If laryngeal reconstruction approaches can ensure VF contact is maintained during phonation (i.e., vertical displacement doesn’t surpass VF medial length), better vocal outcomes are expected.

## 1. Introduction

Acute external laryngeal trauma, which can be either blunt or penetrating, is a potentially life-threatening injury that accounts for 1 in 30, 000 emergency room visits. Blunt force laryngeal trauma is often a result of motor vehicle collisions, sports-related injuries, strangulation, and clothesline-type injuries. Signs of laryngeal trauma vary from patient to patient, which can make it difficult to diagnose. Severe cases have been known to show minimal signs or symptoms before reaching a critical level of obstruction Butler et al. (2005). In virtually all cases, airway evaluation and management are the top priority when diagnosing laryngeal trauma Sniezek & Thomas (2012).

Once the airway is stabilized, further evaluation of the injury is possible. Schaefer’s Classification System is used to categorize the severity of the laryngeal trauma and consists of five groups Schaefer (1992). (1) Minor endolaryngeal hematomas or lacerations without detectable fractures; (2) more severe edemas, hematomas, minor mucosal disruption without exposed cartilage, or nondisplaced fractures; (3) massive edemas, large mucosal lacerations, exposed cartilage, displaced fractures, or vocal fold (VF) immobility; (4) the same as 3, but more severe, with disruption of the anterior larynx, unstable fractures, two or more fracture lines, or severe mucosal injuries; (5) complete laryngotracheal separation. Groups 1 and 2 are often only treated with medicine, humidification, and voice rest, while surgical intervention and a tracheotomy are almost always necessary for treating groups 3-5. Because restoring the airway is the top priority with these injuries, historically, vocal outcomes are a secondary consideration at best during surgical intervention. In fact, vocal outcomes are usually only evaluated post-surgery. Patients are often, but not always, recommended to see a speech pathologist as part of the recovery process. While post-surgical speech therapy is important because it reduces the risk of subsequent phonotrauma arising from vocal compensation Brosch & Johannsen (1999), it cannot correct underlying structural problems. For this reason, it is necessary to determine how morphological changes in the VFs, arising from blunt force trauma, are likely to influence phonation. This information would be helpful in guiding surgical interventions to ensure vocal outcomes are also optimized.

Each blunt laryngeal trauma is unique in how it affects the geometry of the larynx. A review of the literature reveals the most common injuries following blunt laryngeal trauma are (1) displaced laryngeal cartilage fractures, (2) paralysis of at least one VF, and (3) VF scarring Ogura (1975); Kadish et al. (1994); Brosch & Johannsen (1999); Heman-Ackah & Sataloff (2002); Becker et al. (2014). Arytenoid cartilage dislocations and vertical thyroid cartilage fractures are common examples of displaced laryngeal cartilage fractures. Vertical thyroid cartilage fractures are known to cause tears in the thyroarytenoid muscle and ligaments of the VFs, which is why hematomas are often reported Heman-Ackah & Sataloff (2002). Hematomas that are incorrectly diagnosed or ignored can lead to VF scarring. Paralysis occurs due to recurrent laryngeal nerve injuries, which happen in blunt laryngeal trauma when the nerve is either stretched or compressed near the cricoarytenoid joint Sniezek & Thomas (2012). All three of these injuries result in geometric asymmetries between the opposing VFs. Most notably, VF paralysis and displaced cartilage fractures can cause vertical misalignment between the VFs in the inferior-superior direction. Figure 1 depicts an example of this vertical misalignment due to a displaced arytenoid cartilage Ogura (1975).

**Figure 1:**
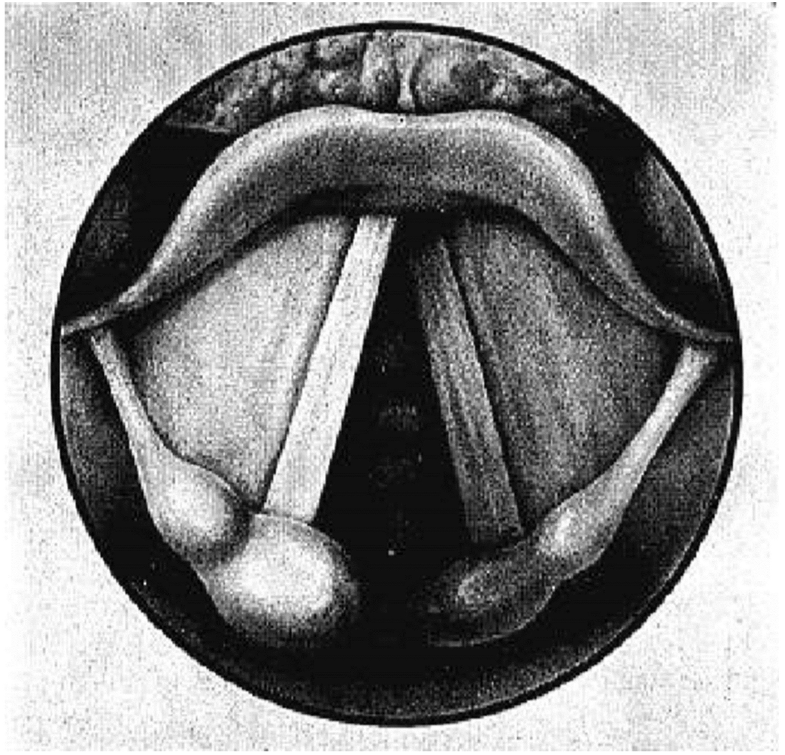
The left arytenoid is superiorly displaced resulting in vertical misalignment between the VFs following blunt force laryngeal trauma. Copyright © 2009, John Wiley and Sons.

There is a large body of prior work that has highlighted how tension asymmetries and VF paralysis influences the biomechanics of phonation, which can be insightful when considering the implications of blunt force trauma. VF asymmetries have been examined using a variety of approaches including reduced-order modeling approaches Steinecke & Herzel (1995); Fraile et al. (2012); Erath et al. (2011); Sommer et al. (2012). numerical simulations Ishizaka & Isshiki (1976); Tao & Jiang (2006), in-vivo canine larynx experiments Sloan et al. (1992), and synthetic modeling of the VFs Pickup & Thomson (2009); Zhang & Hieu Luu (2012), to name a few. This body of work has revealed that asymmetric VF stiffness results in bifurcations Steinecke & Herzel (1995), phase differences Schwarz et al. (2006); Zhang et al. (2013); Lucero et al. (2015); Geng et al. (2016), entrainment of one VF to the dominant VF Zhang (2010); Lucero et al. (2015), subharmonics Mergell et al. (2000), biphonation Tigges et al. (1997), anterior-posterior modes Neubauer et al. (2001); Eysholdt et al. (2003), and irregular chaotic oscillations Titze et al. (1993); Berry et al. (1996); Erath et al. (2011). Despite the insights gained from work related to VF paralysis, there is a surprising lack of investigations that are specifically focused on elucidating how displacement of the laryngeal structure due to cartilage fractures, as most commonly occurs from blunt force trauma, influences vocal outcomes. The sole exception is the recent work of Tokuda and Shimamura Tokuda & Shimamura (2017) who first introduced an inferior-superior level difference between the left and right VFs to study the associated effects on phonation. They measured phonation threshold pressure, fundamental frequency, phase difference, and performed a psychoacoustic experiment to evaluate voice quality, concluding that phonation threshold pressure increased as level difference increased and breathy voice was clearly perceived at inferior-superior level differences between the left and right VFs of 3.3 mm and 6.0 mm, but not at level differences of 1.2 mm or below.

These findings suggest that there is a degree of inferior-superior VF displacement that can be tolerated without severe degradation in voice quality. However, it remains to be determined what this acceptable value is, how it relates to the modified geometry of the VF structure, and how the acoustic, kinematic, and aerodynamic parameters of the VFs are influenced by this vertical VF misalignment. This information would be useful during pre-surgical planning of laryngeal reconstruction to optimize vocal outcomes.

To this end, the objective of this work is to determine an acceptable threshold for vertical VF misalignment that is most likely to lead to acceptable vocal effort, VF vibratory patterns, and vocal outcomes. This is determined by introducing successive amounts of vertical misalignment between synthetic, self-oscillating VF models. Acoustic parameters including frequency, sound pressure level (SPL), jitter, shimmer, and H1-H2 are investigated as a function of vertical misalignment. Kinematic parameters of the VF oscillations are extracted from kymograms obtained using high speed videos to identify amplitude and phase differences in the superior and inferior edges of the left and right VFs. Finally, aerodynamic parameters including the time-averaged flow rate and subglottal pressure are also recorded to provide insight into vocal effort. Section 2 discusses the experimental facility and data collection methods. The results are presented in Section 3, and discussed in Section 4, while Section 5 is left for the conclusions.

## 2. Methods

### 2.1 Flow Facility

The experimental set up consisted of a flow facility representative of the human airway and subglottal tract. A schematic of the flow facility is shown in Figure 2. It was housed inside a WhisperRoom SE2000 acoustic booth with acoustically treated interior walls to minimize acoustic reflections. A Central Pneumatic, 29 Gallon (0.11 m^3^), 2 HP (1.5 kW), 150 psi (1.0 MPa) Cast Iron Vertical Air Compressor provided compressed air that was regulated down to operate at approximately 10 psi (69 kPa). Air flows from the compressor to an in-line Dwyer RMC 103-SSV flow meter. The air exits the flow meter and enters a 0.006 m^3^ cylindrical plenum that represents the lung volume. From the lung plenum, air flows into the subglottal channel, representing the trachea, which has a height and width of 0.022 m. The trachea and lung geometries were prescribed to match the in vivo subglottal acoustic loading. A Dwyer Series 1227 manometer measured the subglottal pressure in this channel. Synthetic, self-oscillating VF models, which are further described in Section 2.2, were mounted to the end of the subglottal channel with 3D printed mounting brackets. A B&K 4189 0.5 in free-field microphone was placed 15 cm away from the superior edge of the VFs at an angle of 45 degrees and recorded the radiated acoustic sound pressure data at a sampling frequency of 80,000 Hz for 0.75 s. A Photron AX200 high speed camera was placed superiorly, 30 cm from the exit plane of the VFs, and captured calibrated high speed video (HSV) at 20, 000 frames *−* per *−* second with a resolution of 640 *×* 480 pixels.

**Figure 2:**
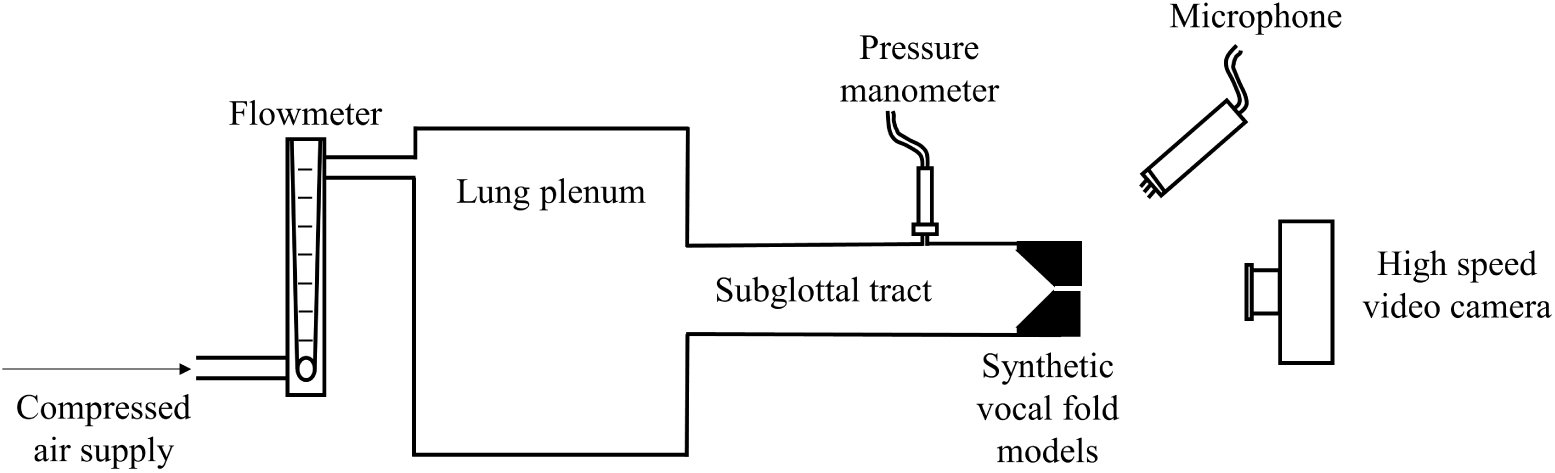
Schematic of the experimental flow facility.

### 2.2 Synthetic, self-oscillating vocal fold models

Synthetic, self-oscillating VF models consisting of four distinct silicone layers were fabricated using a casting method that has been previously employed Murray & Thomson (2012); Motie-Shirazi et al. (2019). Varying ratios of Ecoflex 00-30 (E) and Dragon Skin 10 Fast (D) silicone rubber and Silicone Thinner (Smooth-On, Inc.) were mixed to produce differing values of stiffness for the four layers. The geometry of the VF model, and the corresponding layers, are shown in Figure 3. The layers include (1) the paraglottic space (i.e., adipose tissue), (2) the body layer, (3) the cover layer, and (4) the epithelial layer. The geometry also included an undercut that represents the laryngeal ventricle.

**Figure 3:**
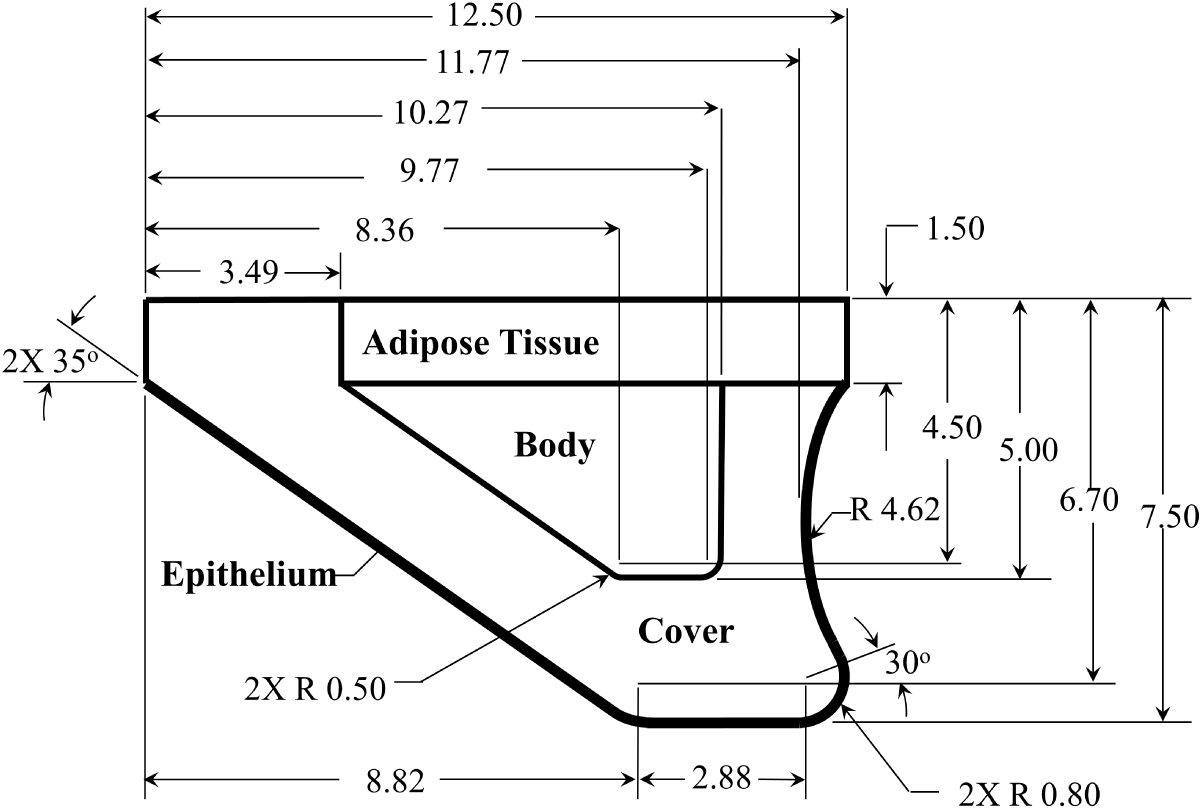
Synthetic VF model geometry. All dimensions are in mm.

The structural properties of each silicone/thinner mixture was measured by casting a core sample of 60.0 mm in diameter that was approximately 1.0 mm thick. A TA Instruments AR 2000 Rheometer performed a frequency sweep at 1% strain from 1 Hz to 100 Hz to determine the elastic and viscous shear moduli. Total shear modulus, *G*^*∗*^, is the sum of elastic and viscous shear moduli. The modulus of elasticity was computed from total shear modulus at 100 Hz as

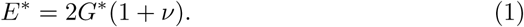

Poisson’s ratio, *ν*, has been measured to be *∼* 0.4 for silicone mixtures at 1% strain rate Stevens (2015).

The ratios of silicone to thinner and the resulting stiffness values for the layers of silicone are reported in Table 1 alongside the range of physiological values. The silicone is a two-part mixture with the type of silicone, and ratio of part A to part B to thinner reported as SiliconePartA:PartB:Thinner, as measured by weight. The corresponding ratios are included in Table 1. As shown, the synthetic VF model material properties were acceptable representations of physiological values.

**Table 1.**
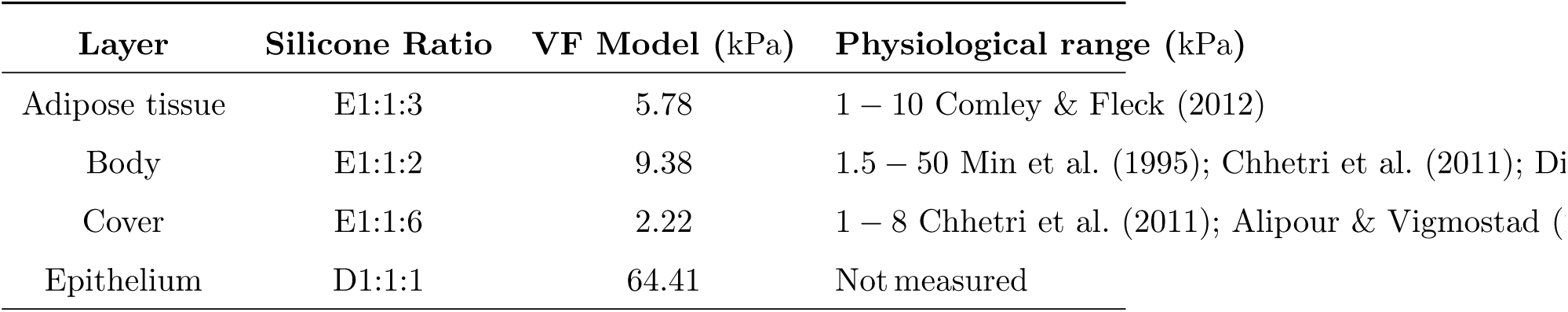
Modulus of elasticity of the four VF model layers resulting from different silicone mixture ratios, with comparison to the physiological values.

| Layer | Silicone Ratio | VF Model (kPa) | Physiological range (kPa) |
| --- | --- | --- | --- |
| Adipose tissue | E1:1:3 | 5.78 | 1 – 10 Comley & Fleck (2012) |
| Body | E1:1:2 | 9.38 | 1.5 – 50 Min et al. (1995); Chhetri et al. (2011); Di |
| Cover | E1:1:6 | 2.22 | 1 – 8 Chhetri et al. (2011); Alipour & Vigmostad ( |
| Epithelium | D1:1:1 | 64.41 | Not measured |

### 2.3 Medial prephonatory pressure

The medial prephonatory pressure (MPP) quantified the amount the opposing medial VF surfaces were compressed against each other when mounted in the flow facility. This was determined independently for each opposing VF by placing the medial surface of each VF model in contact with a flat surface with a flush-mounted Millar Mikro-Cath pressure transducer embedded in it. Each VF was compressed in the medial-lateral direction until the desired pressure was achieved. The distance the model was compressed in the medial direction to achieve the desired MPP, referred to as the medial compressions distance, was then recorded for each VF. Finally, the opposing VFs were mounted in the flow facility, using shims to produce the same medial compression distance that produced the desired MPP (see Motie-Shirazi et al. (2019) for details). For all investigations, a medial prephonatory pressure of 0.6 kPa was used.

### 2.4 Inferior-superior displacement

Inferior-superior vertical displacement was introduced to the upper (i.e., right) VF using shims of 1.0 mm thickness (see Figure 4) that were placed between the VF mounting bracket and the tracheal flow channel, thereby shifting the VF in the superior direction, while maintaining an airtight seal with the flow channel. The amount of displacement in the superior direction was defined by the variable *d*, and is presented as a ratio relative to the inferior-superior length of the medial surface of the VF models expressed as *d/l*_VF_, where *l*_VF_ = 2.88 mm. This is shown in Figure 4. The inferior-superior displacement was increased until the VFs no longer oscillated and therefore ceased producing sound. Data were collected for *d* = 0.0 – 6.0 mm in 1.0 mm increments such that *d/l*_VF_ = 0.0, 0.35, 0.69, 1.04, 1.39, 1.74, and 2.08, respectively. For each inferior-superior displacement the kinematic, aerodynamic, and acoustic measures were collected as a function of subglottal pressure.

**Figure 4:**
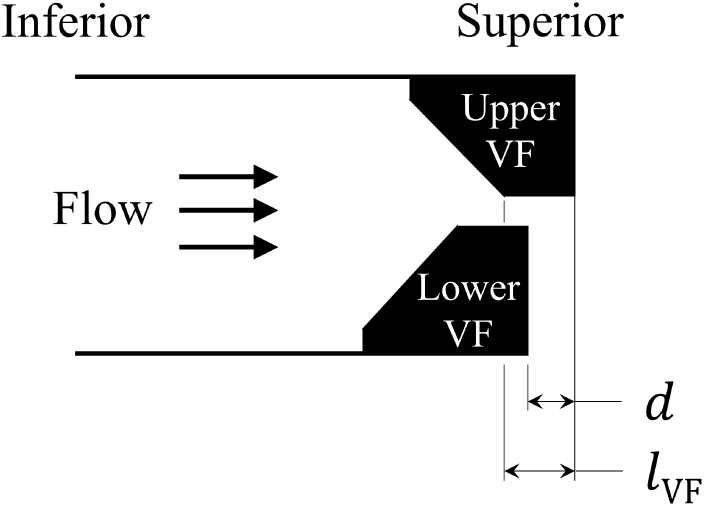
Inferior-superior length of the medial VF surface (*l*_VF_) and vertical displacement, *d*, which was introduced to the upper VF in increments of 1.0 mm.

### 2.5 Aerodynamic, kinematic, and acoustic parameters

Two methods were implemented to determine the impact of the inferior-superior displacement on phonation. First, for each inferior-superior displacement the onset pressure was determined as the subglottal pressure at which self-oscillation occurred while generating a tonal acoustic output. This was found by slowly increasing the subglottal pressure until self-oscillation occurred. Once the onset pressure was determined, the time averaged flow rate, radiated sound pressure level and the VF kinematics were recorded via HSV. The subglottal pressure was then increased in increments of 0.1 kPa until either reaching a value of 1.5 kPa, or when self-oscillation ceased; whichever occurred first. Aerodynamic, kinematic, and acoustic parameters were then compared at the same subglottal pressure for varying VF vertical displacement distances. Next, at each inferior-superior displacement distance, the subglottal pressure was adjusted until the radiated acoustic pressure reached 90 dB. The flow rate, subglottal pressure, and HSV of the kinematics were then recorded, and outputs were compared as a function of vertical misalignment to provide insight into the increased phonation effort needed to overcome the deficiency of VF misalignment and maintain a fixed sound pressure level.

The recorded acoustic pressure was analyzed to compute fundamental frequency, the radiated SPL, jitter, shimmer, and H1-H2. Jitter and shimmer provide measures of cycle-to-cycle variations in the frequency and amplitude of the acoustic signal, respectively. They are computed as

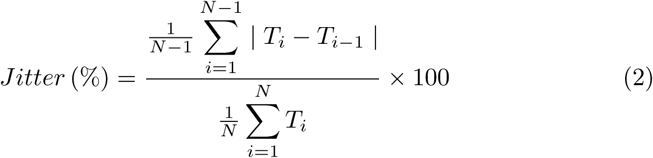

and

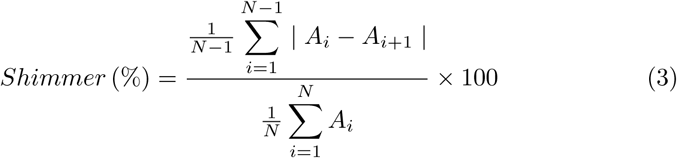

respectively, where *T*_*i*_ is the cycle-to-cycle period of the fundamental frequency, *A*_*i*_ is the cycle-to-cycle amplitude of the acoustic signal, and *N* is the total number of cycles investigated.

Jitter primarily arises due to a lack of vibrational control of the VFs, with pathological voices typically having a higher percentage of jitter Teixeira et al. (2013). Shimmer is correlated with the presence of noise emission and breathiness, where increased values are also usually associated with pathological voice Leclerc et al. (2013). The values of percent jitter and shimmer in normal voice are typically in the range of 0.5 – 1.0 and 0.0 – 3.81, respectively Teixeira et al. (2013); Leclerc et al. (2013).

H1-H2 is the difference between the first and second harmonic peak in the power spectral density plot of an acoustic signal. H1-H2 values are a common measure of perceived voice quality with higher values indicating an increase in open quotient, correlating to an increase in perceived vocal breathiness Kreiman et al. (2012).

The HSV was analyzed to extract kinematic parameters that quantify the amplitude and phase differences between the opposing VFs. Kymograms along the VF midline were extracted from the recorded high-speed videos, and an edge-detection algorithm was used to locate the VF margins in MATLAB. This approach follows the method proposed by Jiang et al.Jiang et al. (2008). In particular, the superior margins during opening, and the inferior margins during closing, were detected for both the upper and lower VFs. This resulted in four sets of data (see Figure 5): upper VF – superior margin (*y*_1_), upper VF – inferior margin (*y*_2_), lower VF – superior margin (*y*_3_) and lower VF – inferior margin (*y*_4_). Each data set was approximated as a Fourier series where the first-order coefficients were determined using least squares regression. The coefficients were then fit to a sine curve for each of the four data sets to quantify the VF motion. This allowed the relative phase shifts between each curve, corresponding to the upper and lower VFs and the inferior and superior margins, of the kymogram to be easily computed. The phase differences were computed between: (1) the superior margin of the upper and lower 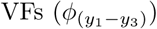, (2) the inferior margin of the upper and lower 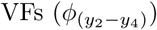, (3) the superior and inferior margins of the upper 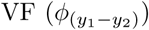, and (4) the superior and inferior margin of the lower 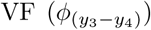. In addition, the oscillation amplitude of the upper and lower VFs were extracted from the kymograms by finding the maximum value for the corresponding VF edges during each cycle, and then calculating the mean over all of the oscillation cycles (*∼* 10 total cycles).

**Figure 5:**
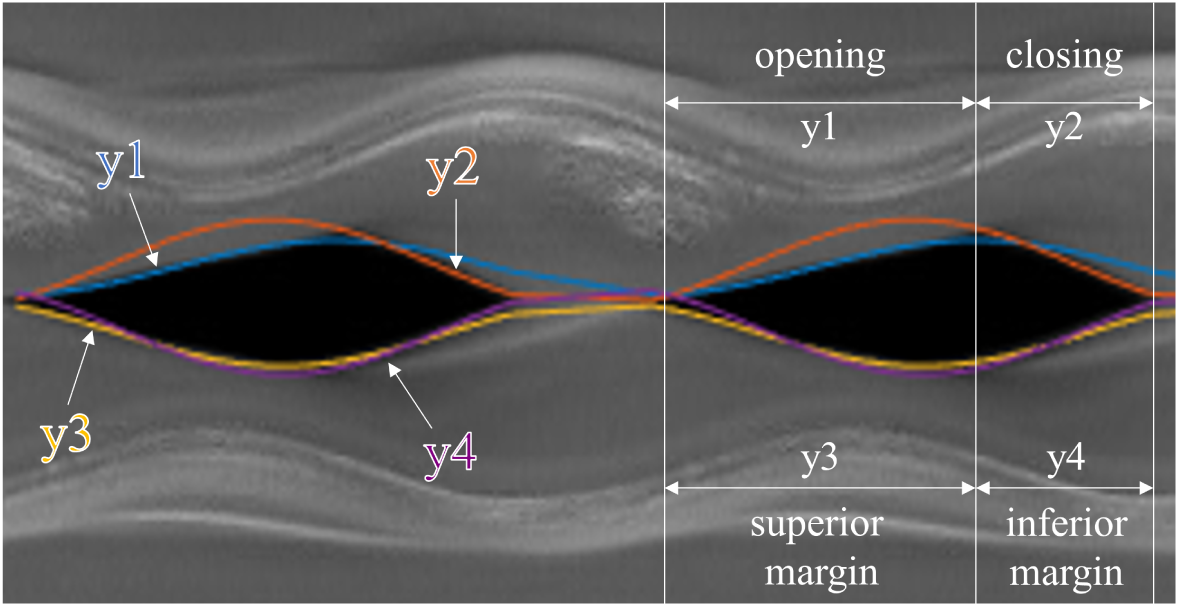
The four curves fitted to the kymogram; *y*_1_ (upper VF – superior margin), *y*_2_ (upper VF – inferior margin), *y*_3_ (lower VF – superior margin), and *y*_4_ (lower VF – inferior margin).

## 3. Results

Data were collected at the seven vertical displacement levels *d/l*_VF_ = 0.0, 0.35, 0.69, 1.04, 1.39, 1.74, and 2.08.

The kymograms for each displacement level are presented in Figure 6 for a subglottal pressure of 1.2 kPa. The corresponding HSV from which the kymograms were extracted are included as supplementary material (Appendix A). For *d/l*_VF_ = 0.0 (symmetric positioning) the kymograms exhibit a clear mucosal wave, with an open quotient (OQ), defined as the time of glottal opening over the period of oscillation, of approximately 0.78. The speed quotient (SQ), defined as the time required for the glottis to transition from closed to maximum opening, divided by the time required for the glottis to transition from maximum opening to closed, is approximately 1.75. These values are within the range of expected physiological observations Kania et al. (2004); Lohscheller et al. (2013). Note, there is a small degree of asymmetry between the upper and lower VFs. This arises due to very slight variations when mounting the VFs in the brackets. Note this asymmetry is imperceptible in the accompanying HSV for *d/l*_VF_ = 0.0 (Appendix A). Even in normal, non-pathological phonation, a small amount of left-right phase asymmetry is not uncommon Mehta et al. (2011). Normal values for left-right phase asymmetry in non-pathological voices can be as high as 0.42 to 1.26 radians Mehta et al. (2011); Sielska-Badurek et al. (2019). At *d/l*_VF_ = 0.0, the phase difference between the superior margin of the upper and lower VFs is 0.52 radians, which is within, or lower than, the expected values for non-pathological phonation.

**Figure 6:**
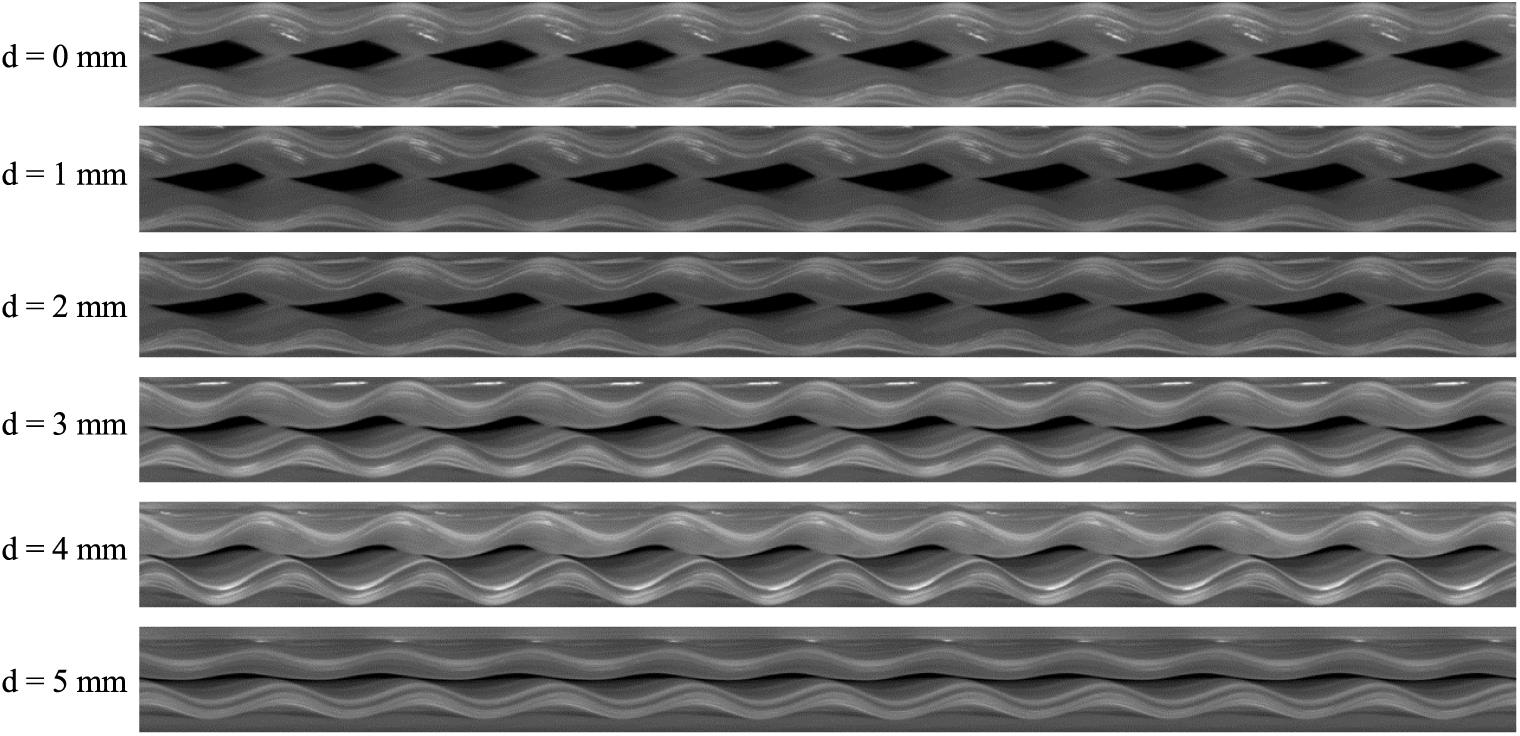
Kymograms consisting of 10 cycles for 6 different levels of *d/l*_VF_ at a subglottal pressure of 1.2 kPa.

As *d/l*_VF_ increased, several trends were evident. The oscillations of the lower VF preceded the upper VF by an increasing amount, resulting in an increase in the absolute value of the phase shift 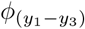. This can be observed by noting the point of maximum opening of the lower VF occurs earlier in the cycle relative to the maximum opening of the upper VF, with increasing values of *d/l*_VF_. Remember, the upper VF is displaced superiorly, relative to the lower VF. In addition both the upper and lower amplitudes decrease as *d/l*_VF_ increases. This can be observed by noting the decrease in visible glottal area as *d/l*_VF_ increases. As *d/l*_VF_ increases, the distinction between the opening phase and closing phase becomes less apparent, indicating a decrease in both 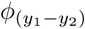 and 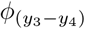. This indicates a decrease in convergent-divergent behavior in the mucosal wave, which is important as the robustness of the mucosal wave is commonly accepted as an indicator of vocal quality.

As shown in Figure 7, 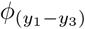 and 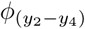 both increased until *d/l*_VF_ = 1.04, then decreased to a value of approximately 0.5 rad, and then increased slightly at the last level. Remember that non-pathological oscillations typically have phase differences in the range of 0.42 and 1.26 radians. Thus, the phase difference between the superior margins becomes pathological when *d/l*_VF_ = 0.69 is reached and the phase difference between the inferior margins becomes pathological when *d/l*_VF_ = 1.04 is reached. As the vertical displacement increased beyond the inferior-superior glottal length (*d/l*_VF_ ≥ 1), both 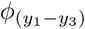 and 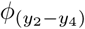 decreased precipitously. As evidenced in the kymogram plots of Figure 6, this occurs as the left-right VFs become entrained

**Figure 7:**
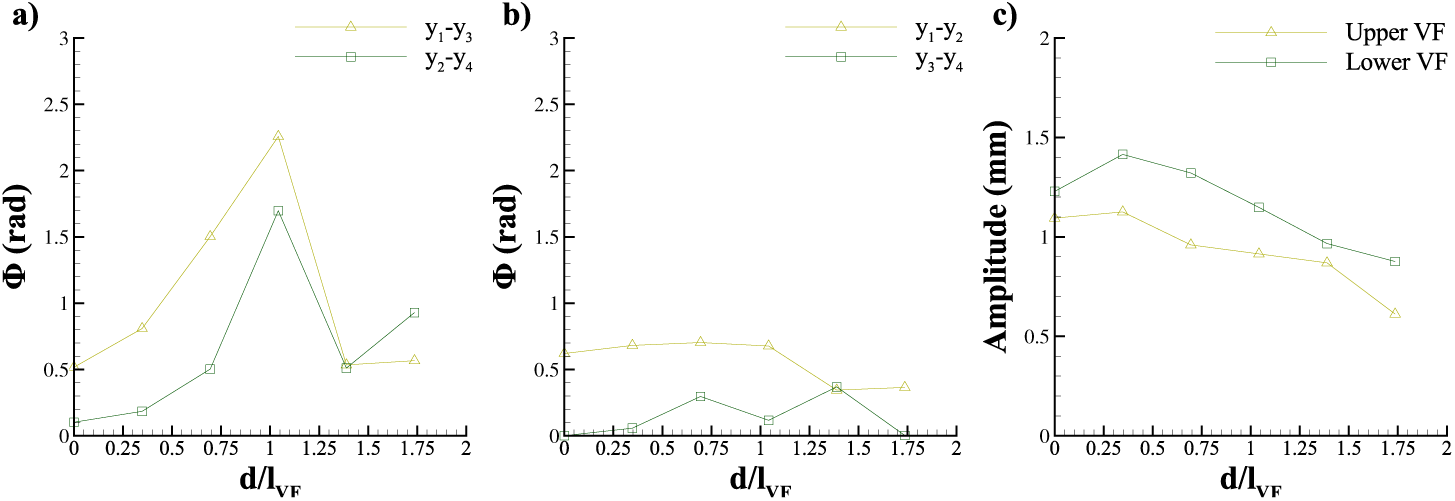
Kinematic variables extracted from the kymograms, plotted as a function of *d/l*_VF_. a) Phase differences between the upper and lower VFs for the superior (*y*_1_ *− y*_3_) and inferior (*y*_2_ *−y*_4_) margins, b) phase differences between the superior and inferior margins for the upper (*y*_1_ *− y*_2_) and lower (*y*_3_ *− y*_4_) VFs, and c) amplitude difference between the upper and lower VF models.

The phase difference between the superior and inferior margins of the same VF did not change as dramatically; 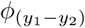 remained relatively constant until *d/l*_VF_ reached 1.04 and then decreased by *∼* 0.3 radians; and 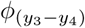 displayed no clear trend. Both the left and right VF amplitudes decreased by *∼* 0.50 mm as *d/l*_VF_ increased.

Figure 8 presents the trends of how flow rate, frequency, SPL, H1-H2, percent shimmer, and percent jitter vary with the inferior-superior displacement, for two subglottal pressures; 1.2kPa and 1.4kPa.

**Figure 8:**
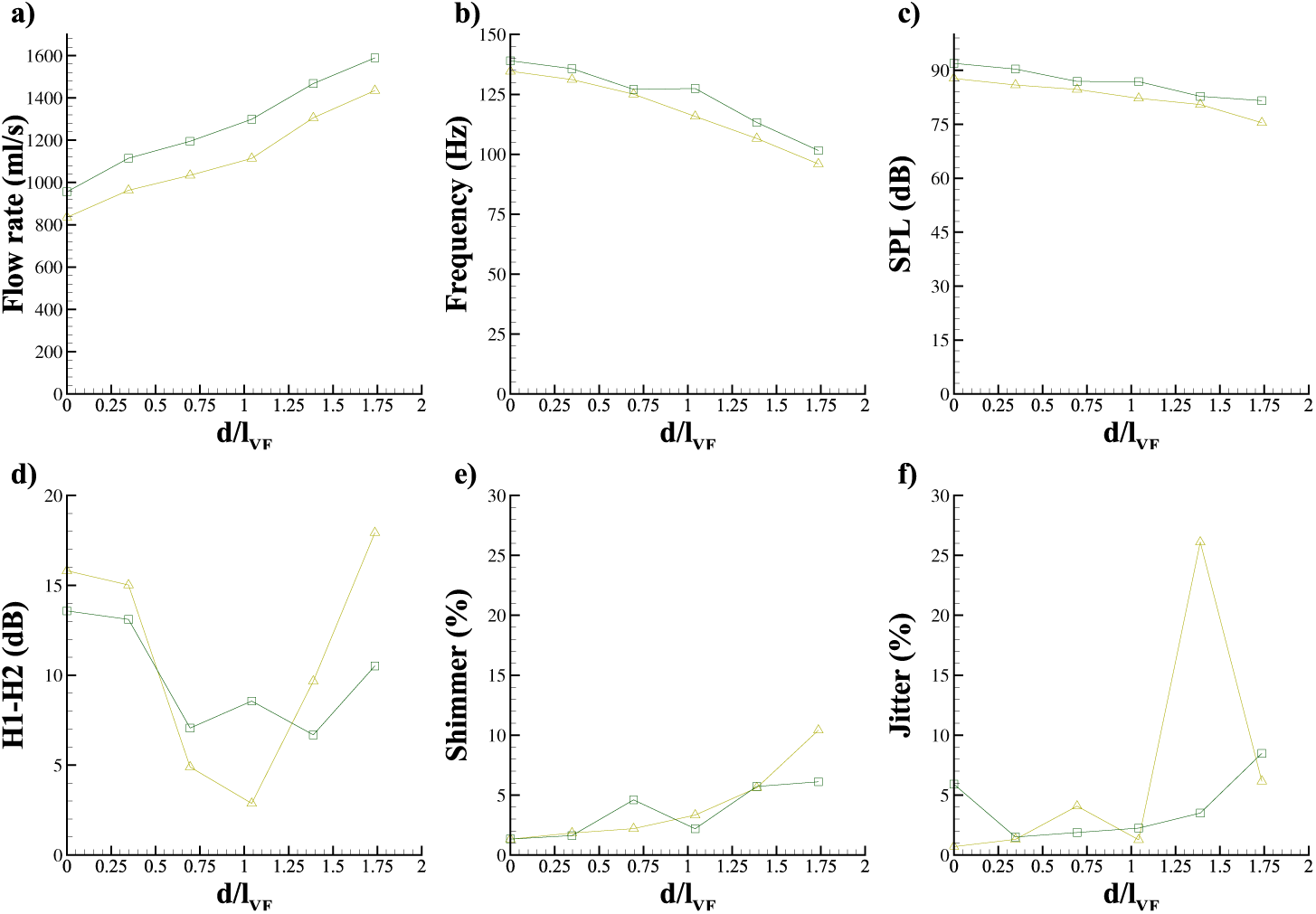
(a) Flow rate, (b) frequency, (c) SPL, (d) H1-H2, (e) shimmer, and (f) jitter, plotted as a function of *d/l*_VF_ at subglottal pressures of 1.2 kPa (delta) and 1.4 kPa (square).

Flow rate, percent shimmer, and percent jitter all increased with increasing vertical displacement. Flow rate increased by 72% at a subglottal pressure of 1.2 kPa and by 67% at a subglottal pressure of 1.4 kPa over a range of *d/l*_VF_ = 0.0 to 1.74. This is consistent with the observation that hoarseness, a hallmark of voice quality in cases of blunt force laryngeal trauma Sniezek & Thomas (2012), is associated with an increase in flow rate. At a subglottal pressure of 1.2 kPa, shimmer steadily increased, reaching a maximum change of 9.1%. At 1.4 kPa, shimmer increased with increasing *d/l*_VF_, except for a slight decrease for the displacement level of 1.04. These results are expected as higher values are typically representative of increased breathiness and pathological voices. Shimmer passes the threshold to detect pathological voices of 3.81 at *d/l*_VF_ = 1.39 for a subglottal pressure of 1.2 kPa, and at *d/l*_VF_ = 0.69 for a subglottal pressure of 1.4 kPa. Jitter generally increased for a subglottal pressure of 1.2 kPa, but decreased for *d/l*_VF_ = 1.04 – 1.39, and then rapidly increased from *d/l*_VF_ = 1.39 – 1.74. At 1.4 kPa, jitter decreased initially, and then steadily increased thereafter. These results are expected as pathological voice is associated with higher values of percent jitter. Jitter passes the threshold to detect pathological voices of 1.04 at *d/l*_VF_ = 0.35 for subglottal pressures of 1.2 kPa and 1.4 kPa. Jitter becomes pathological as soon as inferior-superior displacement is introduced. Shimmer better tolerates the introduction of the displacement and doesn’t become pathological until a displacement of at least 69% of the VF medial length has been reached.

Frequency and SPL both decreased with increasing vertical displacement. Frequency decreased by 20% and 27% for subglottal pressures of 1.2 kPa and 1.4 kPa, respectively. SPL decreased by 14% and 11% at subglottal pressures of 1.2 kPa and 1.4 kPa, respectively. For both subglottal pressures, SPL decreased by at least 10 dB, which corresponds to a two-fold decrease in volume or loudness.

For a subglottal pressure of 1.2 kPa, H1-H2 decreased until *d/l*_VF_ = 1.39 and then subsequently increased. For a subglottal pressure of 1.4 kPa, H1-H2 followed a similar trend except for an increase from *d/l*_VF_ = 1.04 – 1.39 and a decrease from *d/l*_VF_ = 1.39 – 1.74. These results are somewhat surprising because higher H1-H2 values are correlated to an increase in perceived breathiness. The increase after *d/l*_VF_ = 1.04 is consistent with prior understanding, but the initial decrease was unexpected.

To determine the effect of compensating for reduced SPL due to vertical VF misalignment, the kinematic, aerodynamic, and acoustic parameters were computed by adjusting the subglottal pressure until a target SPL level of 90 dB was achieved. This value was selected based on the ability to achieve it over the broadest range of inferior-superior displacement levels, while also achieving self-sustained VF oscillations. Figure 9 shows how the flow rate, frequency, subglottal pressure, H1-H2, shimmer, and jitter vary as a function of vertical displacement for a constant SPL output of 90 dB at each displacement level.

**Figure 9:**
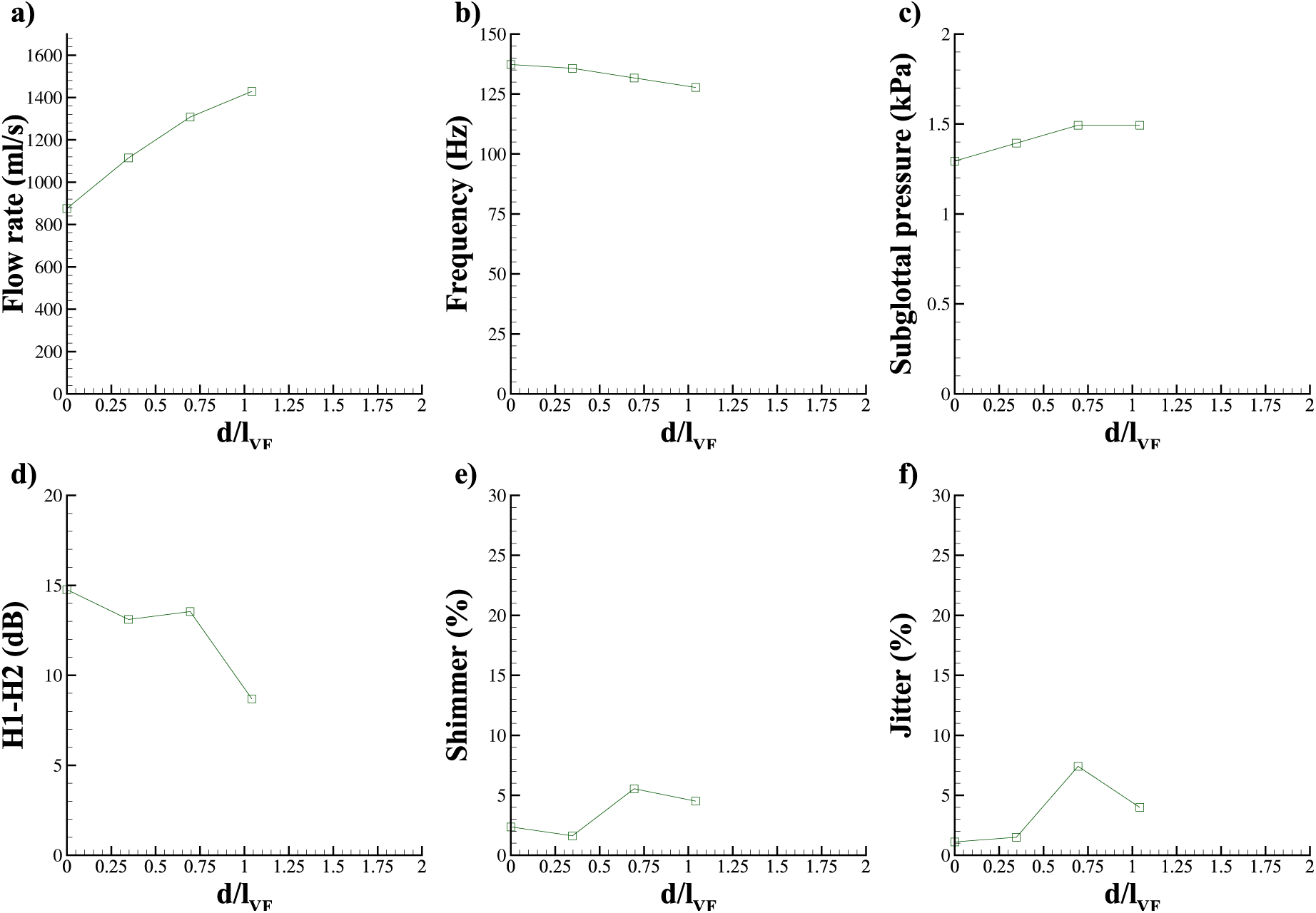
(a) Flow rate, (b) frequency, (c) subglottal pressure, (d) H1-H2, (e) shimmer, and (f) jitter, plotted as a function of *d/l*_VF_ at a constant SPL of 90 dB.

The frequency decreases and flow rate increases with increasing vertical displacement, consistent with the trends in Figure 8. Subglottal pressure steadily increases until *d/l*_VF_ = 0.69 and then remains constant. The observed trends in flow rate and subglottal pressure are consistent with increased effort required to achieve the desired SPL. H1-H2, shimmer, and jitter also demonstrate trends consistent with the plots in Figure 8. For values of *d/l*_VF_ greater than 1.04, the target SPL level is no longer able to be achieved.

## 4. Discussion

It is interesting to note that the more superior VF (the upper VF in the kymograms of Figure 6 continues to experience more robust oscillations with a largely constant phase shift of 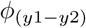, while the lower VF experiences a continuously increasing phase shift (see also, Figure 7). This seems to indicate that from an aerodynamic standpoint, the more inferior VF does not experience the same energy capture from the fluid that the upper VF does.

A key finding is that when *d/l*_VF_ ≳ 1.0, the aerodynamic, kinematic, and acoustic voice measures exhibit profound changes that are largely pathological in nature. Most evident is that after *d/l*_VF_ surpasses *∼* 1.0, the target SPL is no longer able to be produced. This is a clear indicator that voice production has become significantly quieter. The increase in flow rate and subglottal pressure indicates there is a significant increase in the effort required to phonate. The corresponding increases in phase difference between the left and right VFs suggest less efficient sound production due to the onset of asymmetric, irregular VF vibration. Once *d/l*_VF_ reaches values greater than *∼* 1.0, jitter and shimmer values have already surpassed the threshold value used to identify pathological voices. It should be mentioned, however, that the precise value of these measures may also be influenced by the modeling approach. Nevertheless, the trend of worsening outcomes as a function of increasing *d/l*_VF_, with the most pronounced effect occurring when *d/l*_VF_ *>* 1.04, is evident.

For values of *d/l*_VF_ less than 1.0, the aerodynamic, kinematic and acoustic voice measures are still worsening with increasing offset distance, but the change as a function of offset distance is much less pronounced than when *d/l*_VF_ *>* 1.04. This suggests that as long as the vertical misalignment is less than the inferior-superior length of the medial VF surface it is tolerated by the VFs, as evidenced by the kinematic, aerodynamic, and acoustic outcomes. That is to say, as long as the medial surfaces are still aligned such that they can achieve contact, and closure, vocal outcomes are not as likely to experience severe degradation. This is consistent with the findings of Tokuda and Shimamura Tokuda & Shimamura (2017) who, while not explicitly investigating this relationship, identified that breathy voice was perceived when *d/l*_VF_ *>* 1.0, but not perceived when *d/l*_VF_ *<* 1.0.

This is a significant finding that has clear implications for surgical intervention. During laryngeal reconstruction, care should be taken to ensure vertical alignment is preserved, as misalignments greater than the inferior-superior length of the medial VF surface have been shown in this work to have drastic consequences on phonation outcomes. It should, however, be emphasized that inferior-superior displacement is only one measure of the structural VF changes that may arise from blunt force laryngeal trauma. Consequently, there are likely to be other measures of VF structural integrity that will also influence vocal outcomes. Nevertheless, this work provides clear insight into the importance of vertical alignment, and establishes a foundation upon which future investigations can be pursued.

## 5. Conclusions and future research

Inferior-superior displacement between the left and right VFs was investigated to determine if there was an acceptable level of vertical misalignment the VFs could tolerate before voice outcomes became pathological. This work is directly applicable to surgical remediation following blunt force laryngeal trauma. Synthetic, self-oscillating VF models were utilized for the investigations. Inferior-superior displacement was modeled by introducing increasing vertical displacement distances in increments of 1.0 mm. It was determined that if the inferior-superior VF displacement distance surpassed the length of the inferior-superior VF medial length, voice production degraded significantly, becoming pathological. If laryngeal reconstruction approaches can ensure VF contact is maintained during phonation (i.e., vertical displacement doesn’t surpass VF medial length), better vocal outcomes are expected. Future research should aim to provide a more encompassing view of blunt force laryngeal trauma by introducing VF stiffness asymmetries, as occurs due to VF scarring and paralysis, as well as considering angular misalignment of the opposing VFs due to arytenoid cartilage displacement.

## Data Availability

I collected the data myself

## 6. Conflict of interest

There is no conflict of interest with the authors and any aspect of the present work.

## 7. Appendix A: Supplementary material

Supplementary material associated with this article can be found in the online version at

## Acknowledgements

This work was funded in part by the National Institutes of Health (NIH) National Institute on Deafness and other Communication Disorders Grant P50 DC015446.

## Notes

### Competing Interest Statement

The authors have declared no competing interest.

### Author Declarations

No IRB/oversight body approval or exemption was needed

